# In-Hospital ACEI Use and Long-Term Prognosis in Discharged Type 2 Myocardial Infarction Patients

**DOI:** 10.1101/2025.10.09.25337699

**Authors:** Jing-yin Zhang, Jia-ni Liu, Wei-ye Feng, Yun-yue Luo, Wu-lin Li, Yue-Li, Yu-xin Wang, Xiao-ya Ma, Xue-feng Ju, Fei Wang

**Author notes:** **Co-authors:** Jing-yin Zhang, Jia-ni Liu and Wei-ye Feng contributed equally to this work. **Correspondence:** Fei Wang, Department of Emergency and Critical Care Medicine, No.1, Chengbei Rd, Jiading District, Shanghai, China., Xue-feng Ju, Emergency Physician’s Office, No.1, Chengbei Rd, Jiading District, Shanghai, China.

## Abstract

**Objective:** To explore the relationship between angiotensin-converting enzyme inhibitor (ACEI) use during hospitalization and both short-term and long-term prognosis of type 2 myocardial infarction (T2MI) patients after discharge.

**Methods:** This was a retrospective cohort study based on Medical Information Mart for Intensive Care IV(MIMIC-IV). Adult critically ill patients with T2MI were included in the analysis. The exposure was ACEI use during ICU stay. The primary outcome measure was 3-year all-cause mortality. Propensity score matching (PSM) was performed at a 1:1 ratio. Multivariate analysis was used to adjust for confounding factors.

**Results:** A total of 1086 T2MI patients were included. The PSM cohort included 590 patients, with 295 patients in each group. ACEI use significantly reduced the 3-year all-cause mortality (hazard ratio [HR], 0.55; 95% confidence interval [CI], 0.41-0.74; P<0.001). Both univariate and multivariate Cox proportional hazards analyses indicated that ACEI use reduced the risk of 3-year all-cause mortality (univariate HR, 0.55; multivariate HR, 0.59; all P<0.001). Sensitivity analysis using the entire cohort also showed that ACEI use significantly reduced 3-year all-cause mortality (HR, 0.58; 95% CI, 0.42-0.80; P<0.001). In addition, ACEI use significantly reduced 180-day all-cause mortality (HR, 0.43; 95% CI, 0.29-0.64; P<0.001). Both univariate Cox proportional hazards model analysis (HR, 0.43; 95% CI, 0.29-0.65; P<0.001) and multivariate Cox proportional hazards model analysis (HR, 0.49; 95% CI, 0.33-0.72; P<0.001) indicated that ACEI use reduced the risk of 180-day all-cause death in patients. The study results also showed that ACEI use had no effect on the risk of 3-year all-cause readmission (HR, 1.003; 95% CI, 0.776-1.295; P=0.984) and 3-year T2MI recurrence risk (HR, 1.14; 95% CI, 0.64-2.05; P=0.654).

**Conclusion:** ACEI use significantly reduces both short-term and long-term all-cause mortality in discharged T2MI patients, while it does not affect patients’ readmission rates or T2MI recurrence rates. These findings expand the clinical applications of ACEI in populations with myocardial infarction of different pathogenesis. Further prospective studies are needed to determine the optimal administration strategy of ACEI.

**Key Question:** Does ACEI use during hospitalization affect the long-term prognosis of T2MI patients after discharge?

**Key Finding:** In discharged T2MI patients, ACEI use significantly reduced the 3-year and 180-day all-cause mortality, but had no effect on the risk of 3-year all-cause readmission or T2MI recurrence.

**Take Home Message:** ACEI use significantly reduces both short-term and long-term all-cause mortality in discharged T2MI patients; nevertheless, it does not affect readmission or T2MI recurrence rates. These findings broaden the clinical use of ACEI to include patients with myocardial infarction caused by different pathophysiological mechanisms.

## Introduction

Type 2 myocardial infarction (T2MI) is myocardial necrosis caused by an imbalance between myocardial oxygen supply and demand. This imbalance is not related to coronary plaque rupture [1]. Its main risk factors include reduced myocardial oxygen supply, such as anemia, hypoxemia, and coronary artery spasm, or increased myocardial oxygen demand, such as tachycardia, hypertensive crisis, and severe infection [1, 2]. These risk factors are more common in critically ill patients. The treatment strategy for T2MI mainly targets the primary disease, such as correcting anemia, improving hypoxemia, or managing shock, rather than coronary revascularization [3]. Existing studies have found that, in addition to treating the primary disease, lipid-lowering therapy (hazard ratio [HR] 0.77, 95% confidence interval [CI] 0.61-0.97) [4], sodium-dependent glucose transporters 2(SGLT2) inhibitors (HR 0.67, 95% CI 0.41-1.10) [5, 6], and glucagon-like peptide-1(GLP-1) agonists (HR 0.65, 95% CI 0.46-0.92) [7] can significantly reduce the incidence of T2MI. However, in patients diagnosed with T2MI, it remains unclear whether antiplatelet agents [8–10], lipid-lowering drugs [8, 11], beta-blockers [8–10], and ACEI [8, 10] provide clinical benefits. The treatment of T2MI still lacks standardized plans or recommendations to date [12–14].

It is worth noting that in patients with type 1 myocardial infarction (T1MI), ACEI mainly exert their effects by inhibiting the renin-angiotensin system (RAS). This inhibition reduces the production of angiotensin II (Ang II)and increases bradykinin levels. These actions lead to vasodilation, reduction of cardiac load, and delaying myocardial remodeling, thereby lowering the mortality rate of patients with acute myocardial infarction(AMI) [15–17]. Additionally, relevant guidelines clearly state that for myocardial infarction patients with a left ventricular ejection fraction (LVEF) ≤ 40%, or those complicated with hypertension, diabetes mellitus or chronic kidney disease, ACEI is recommended to be administered as early as possible and continued over the long term in the absence of contraindications [18]. Therefore, these findings indicate that the use of ACEI is of great significance in the T1MI population. A recent study found that ACEI use may reduce 2-year mortality among T2MI patients, suggesting that ACEI benefits not only the T1MI population, traditionally affected by coronary plaque rupture, but also potentially offers protection to T2MI patients.

However, during the acute phase of the disease, the severity of the primary condition and treatment strategy may have a stronger impact on short-term mortality risk in T2MI patients. This influence may reduce the prognostic value of traditional myocardial infarction treatments, such as the use of ACEI. These findings suggest that T2MI patients with improvement in their primary condition after treatment require increased clinical attention. Therefore, this study aims to explore the relationship between ACEI use during hospitalization and the short- and long-term prognosis of T2MI survivors, providing a basis for ACEI application in these patients.

## Methods

### Data sources

All data were obtained from the Medical Information Mart for Intensive Care IV version 3.1 (MIMIC-IV 3.1) database [20]. This openly accessible critical care database contains electronic medical records of 546,028 patients. The data were collected by Beth Israel Deaconess Medical Center in Boston, Massachusetts, from 2008 to 2022. Patients’ demographic information, laboratory tests, vital signs, hospital status, medication, and surgical procedures are documented in detail in the MIMIC-IV database. All information about patients’ identification has been anonymized, and all identifiable information has been removed. Therefore, informed consent was waived. The author completed the required data research training from the Collaborative Institutional Training Initiative to obtain database access permission (Record ID: 52,310,626). Our study follows the TRIPOD guideline statement for transparent reporting of multivariable prediction models for individual prognosis or diagnosis [21].

### Study population

In the MIMIC-IV 3.1 database, all patients with a first admission from 2015 to 2022 were extracted. The diagnosis of T2MI was determined by the international classification of diseases(ICD)10 code I12.A1.

#### Inclusion criteria

Patients with the ICD10 diagnostic code I12.A1 were included.

#### Exclusion criteria

1. Patients who died during hospitalization.
2. Patients with missing data on ACEI medication.

### Data collection

Data extraction was performed using Structured Query Language (SQL) scripts retrieved from the GitHub website (https://github.com/MIT-LCP/mimic-iv).

Relevant patient information was collected, including the following categories:

**Demographic data:** gender and age;

**Comorbidities:** Charlson Comorbidity Index (CCI), old myocardial infarction, atrial fibrillation, congestive heart failure, cerebrovascular disease, chronic lung disease, diabetes mellitus, kidney disease, malignant tumor, hypertension, and hyperlipidemia;

**Risk factors for T2MI**: sepsis, acute respiratory distress syndrome(ARDS), acute kidney injury, septic shock, tachyarrhythmia, bradyarrhythmia, acute respiratory failure with hypoxemia, hypotension, and acute hemorrhagic anemia;

**Medication use:** vasoactive drugs, anticoagulants, antiplatelet drugs, statins, diuretics, angiotensin II receptor blockers (ARBs), digitalis drugs, antibiotics, and beta-blockers.

**Special treatments received during hospitalization**: continuous renal replacement therapy (CRRT), mechanical ventilation(MV), and length of hospital stay.

### Exposure and outcome

Exposure was defined as the use of ACEI at any time during the ICU stay. Data on ACEI exposure were obtained from medical prescription records. Patients with missing data on ACEI exposure were excluded from the analysis.

The primary endpoint was 3-year all-cause mortality.

Secondary endpoints included 180-day all-cause mortality, 3-year readmission rate, and T2MI recurrence rate.

### Statistical analysis

As a retrospective analysis, there was no predefined statistical analysis plan. No statistical power calculation was performed, and the sample size was based on the available data in the database. The study cohort was divided into two groups: patients who received ACEI treatment (ACEI use group) and those who did not (non-use group). Variance inflation factors(VIF) are used to detect multicollinearity among variables.

For continuous data, the Shapiro-Wilk test was used to assess normality. If the data were normally distributed, they were expressed as mean ± standard deviation, and inter-group comparisons were performed using the independent samples t-test. If not normally distributed, they were presented as median (interquartile range [IQR]), and inter-group comparison was conducted with the Mann-Whitney U test. Categorical data were expressed as percentages, and inter-group comparison was made using the chi-square test. To estimate the HR and its 95% CI for the primary outcome (3-year all-cause mortality), we used a Cox proportional hazards model to assess the independent effect of the exposure factor on survival time. The cumulative incidence of 3-year all-cause mortality was estimated using the Kaplan-Meier method and compared by the log-rank test. For binary outcomes, including 180-day all-cause mortality and 180-day and 3-year all-cause readmission rates, Cox regression calculated the HR and its 95% CI. The cumulative incidences of secondary outcomes were also estimated by the Kaplan-Meier method to facilitate result interpretation. All data were analyzed using R software (version 4.3.3), and a two-tailed P-value <0.05 was considered statistically significant.

**PSM**The main analysis was conducted in the matched cohort to explore the association between ACEI use and both primary and secondary outcomes. We used PSM to adjust for variables. The probability of each patient receiving ACEI treatment (i.e., propensity score) was calculated using a logistic regression model. The selection of variables for the propensity score model used for matching was based on published consensus statements in the literature, including gender, age, CCI, old myocardial infarction, atrial fibrillation, congestive heart failure, cerebrovascular disease, chronic lung disease, diabetes mellitus, nephropathy, malignancy, hypertension, hyperlipidemia, sepsis, ARDS, acute kidney injury, septic shock, tachyarrhythmia, bradyarrhythmia, acute respiratory failure with hypoxemia, hypotension, acute hemorrhagic anemia, vasoactive agents, anticoagulants, antiplatelet agents, statins, diuretics,ARBsdigitalis, antibiotics, beta-blockers,CRRT MV, and length of hospital stay. Matching was performed at a 1:1 ratio using the nearest neighbor method, with a caliper width set at 0.05, and no repeat matching. The balance of variables between groups before and after matching was assessed by the standardized mean difference (SMD), and an SMD value less than 0.10 was considered to indicate balance achieved. In the matched cohort dataset, variables with P < 0.05 in the univariate analysis—such as CCI, old myocardial infarction, congestive heart failure, malignancy, hyperlipidemia, antiplatelet drugs, statins, diuretics,ARBsand beta-blockers—were included in the multivariate analysis for adjustment.

### Subgroup analysis

Subgroup analyses in the matched cohort were based on age groups (≤65 years vs 65-80 years vs ≥80 years), presence of tachycardia (No vs Yes),CCIcategories (<7, 7-8, >8), use of digitalis drugs (No vs Yes), and length of hospital stay (≤6, 6-12, and ≥12 days).

### Sensitivity analysis

We performed a sensitivity analysis on the entire dataset to test the robustness of the results obtained in the matched cohort. The variables involved included demographic factors (gender, age), clinical indices (CCI), comorbidities (old myocardial infarction, atrial fibrillation, congestive heart failure, cerebrovascular disease, chronic lung disease, diabetes mellitus, nephropathy, malignancy, hypertension, hyperlipidemia, sepsis, ARDS, acute kidney injury, septic shock, tachyarrhythmia, bradyarrhythmia, acute respiratory failure with hypoxemia, hypotension, acute hemorrhagic anemia), medications (vasoactive drugs, anticoagulants, antiplatelet drugs, statins, diuretics,ARBsdigitalis, antibiotics, beta-blockers), and treatments (CRRT, MV), as well as length of hospital stay.

## Results

### Patient selection

A total of 122,905 records were identified. After applying the inclusion and exclusion criteria, 1,086 patients with T2MI were finally included, of whom 302 (27.8%) received ACEI therapy during hospitalization. The matched cohort consisted of 590 patients, with 295 in each group. Figure 1 illustrates the patient selection process.

**Figure 1.**
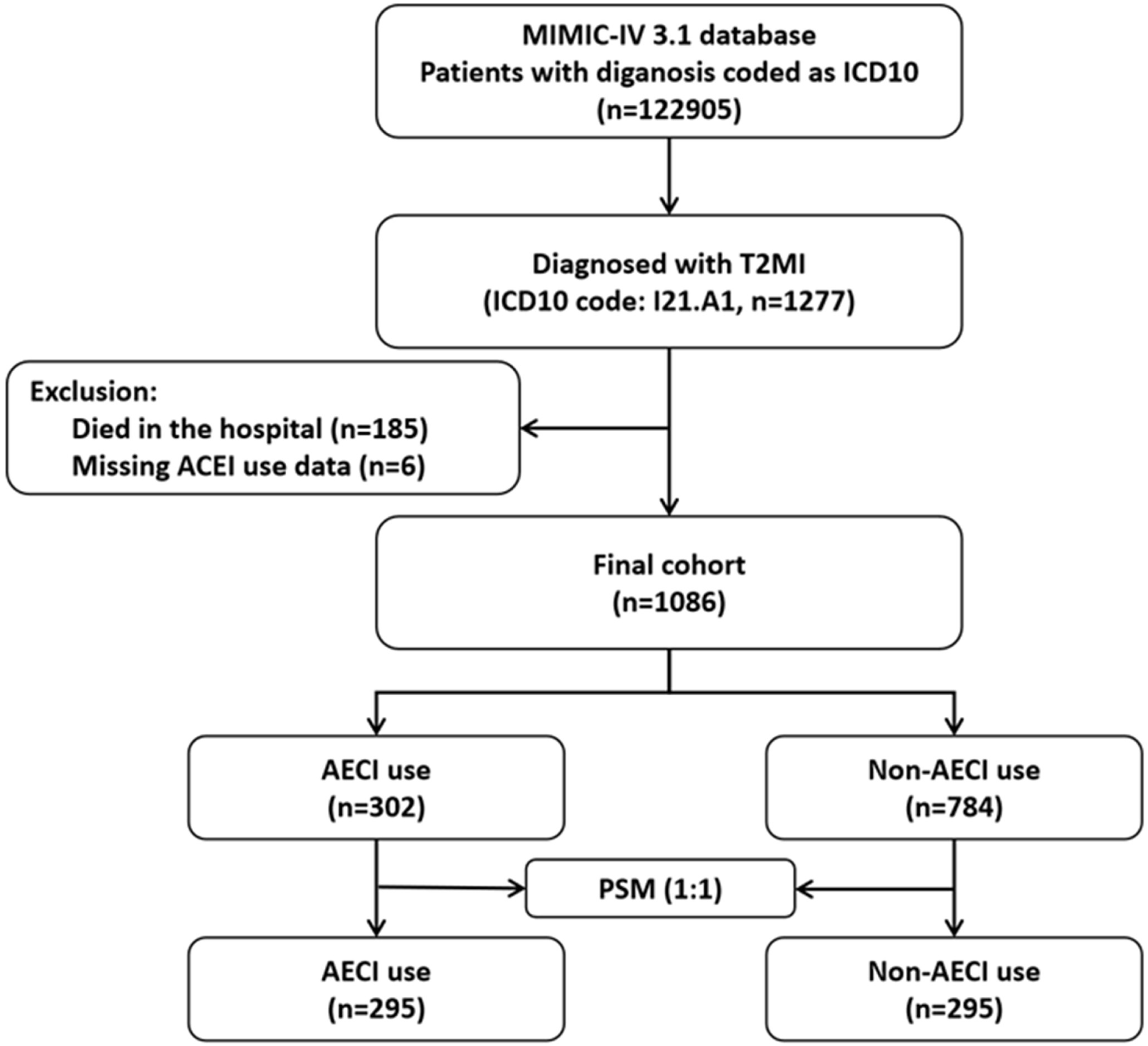
Flowchart of study population

### Cohort characteristics

Table 1 presents the baseline characteristics of patients before and after matching. The VIF of each variable was less than 5, indicating no multicollinearity (Supplementary Table 1). In the entire cohort, significant differences were observed in confounding factors—including CCI, old myocardial infarction, congestive heart failure, malignant tumor, and hyperlipidemia—as well as medication use such as antiplatelet, statins, diuretics, ARBs and beta-blockers between patients who received ACEI treatment (P < 0.05) (Supplementary Table 2). Matching improved variable balance, achieving an absolute SMD of less than 0.10, as shown in Supplementary Figures 1 and 2, which illustrate the distribution balance before and after PSM.

**Table 1.**
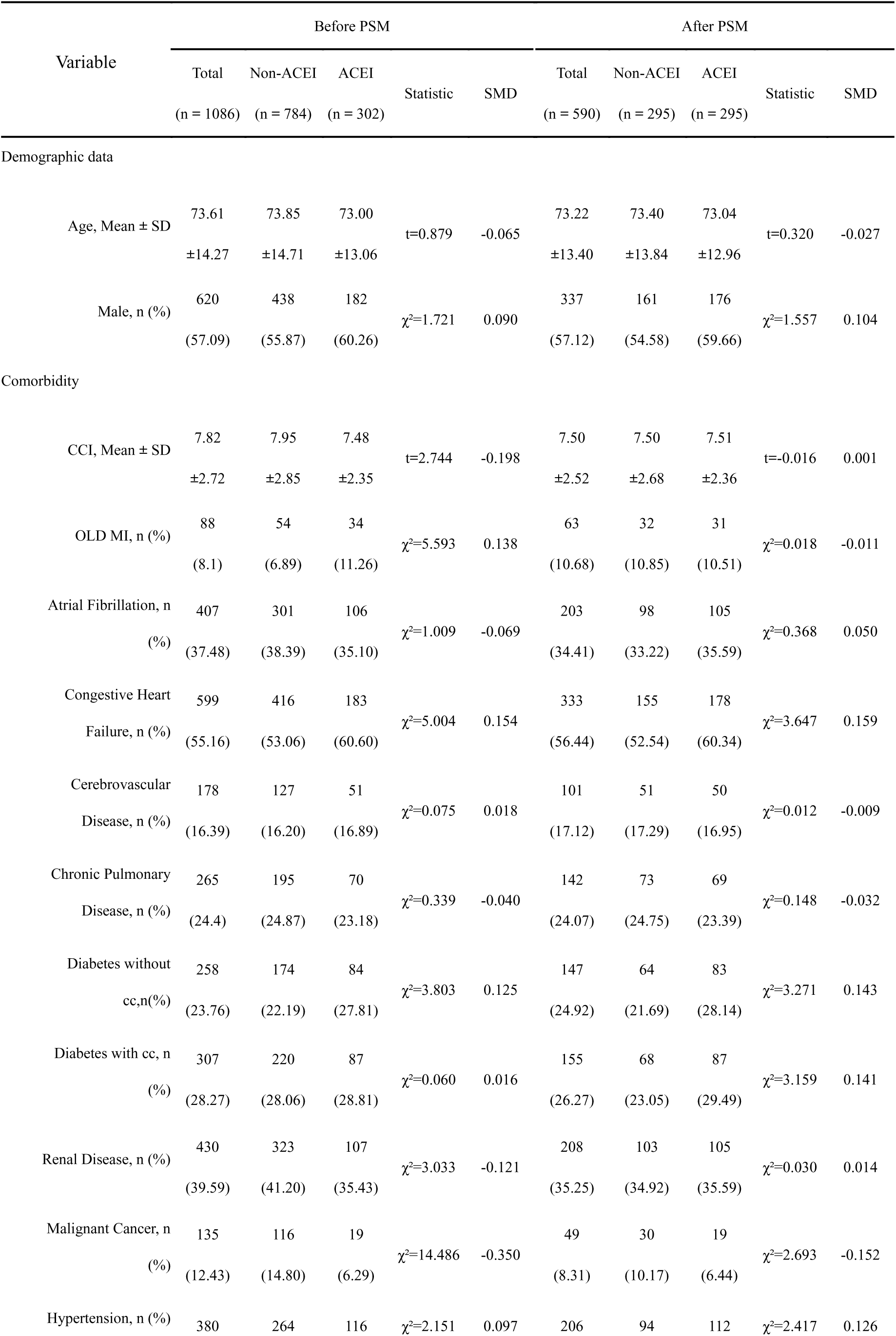
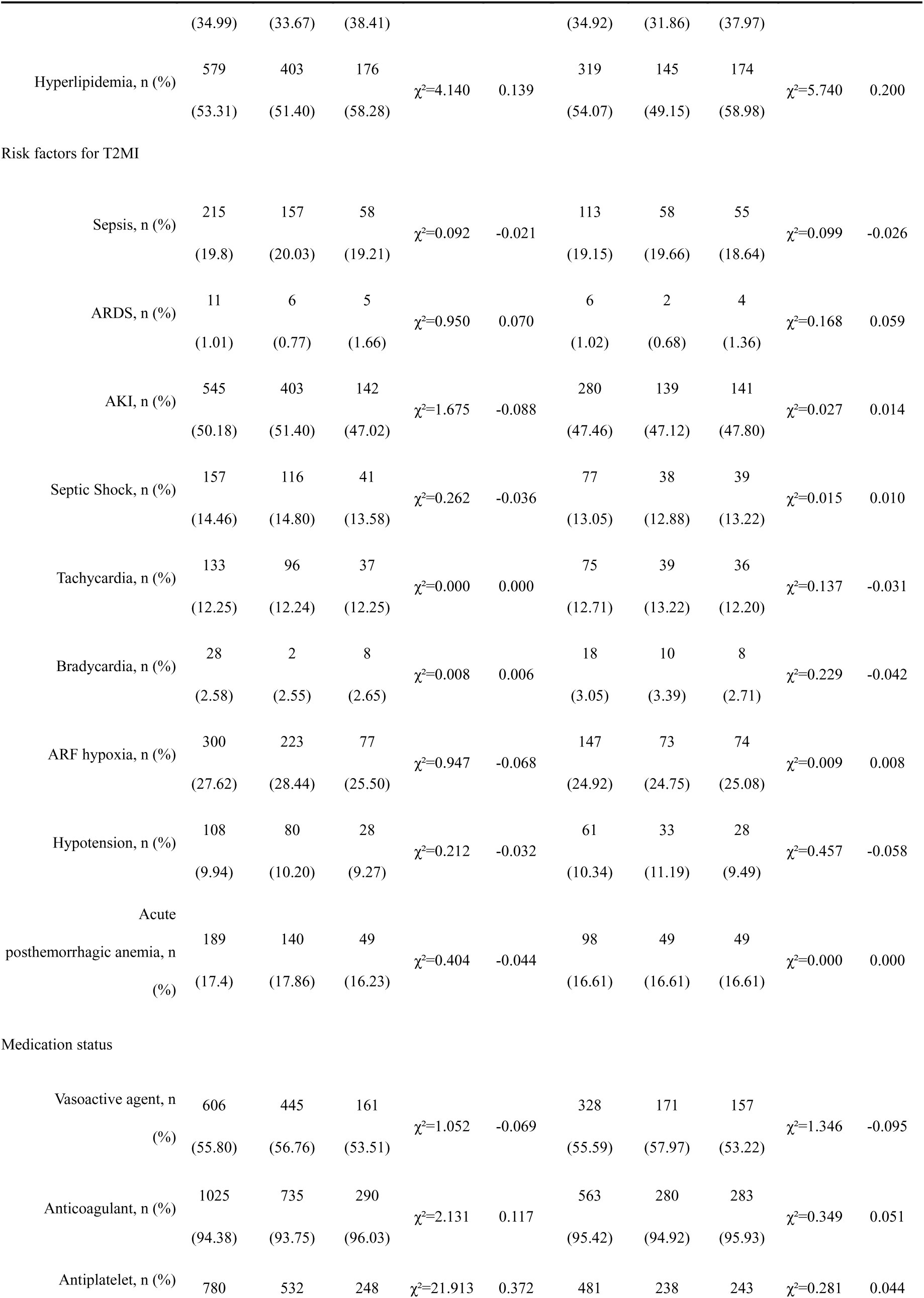
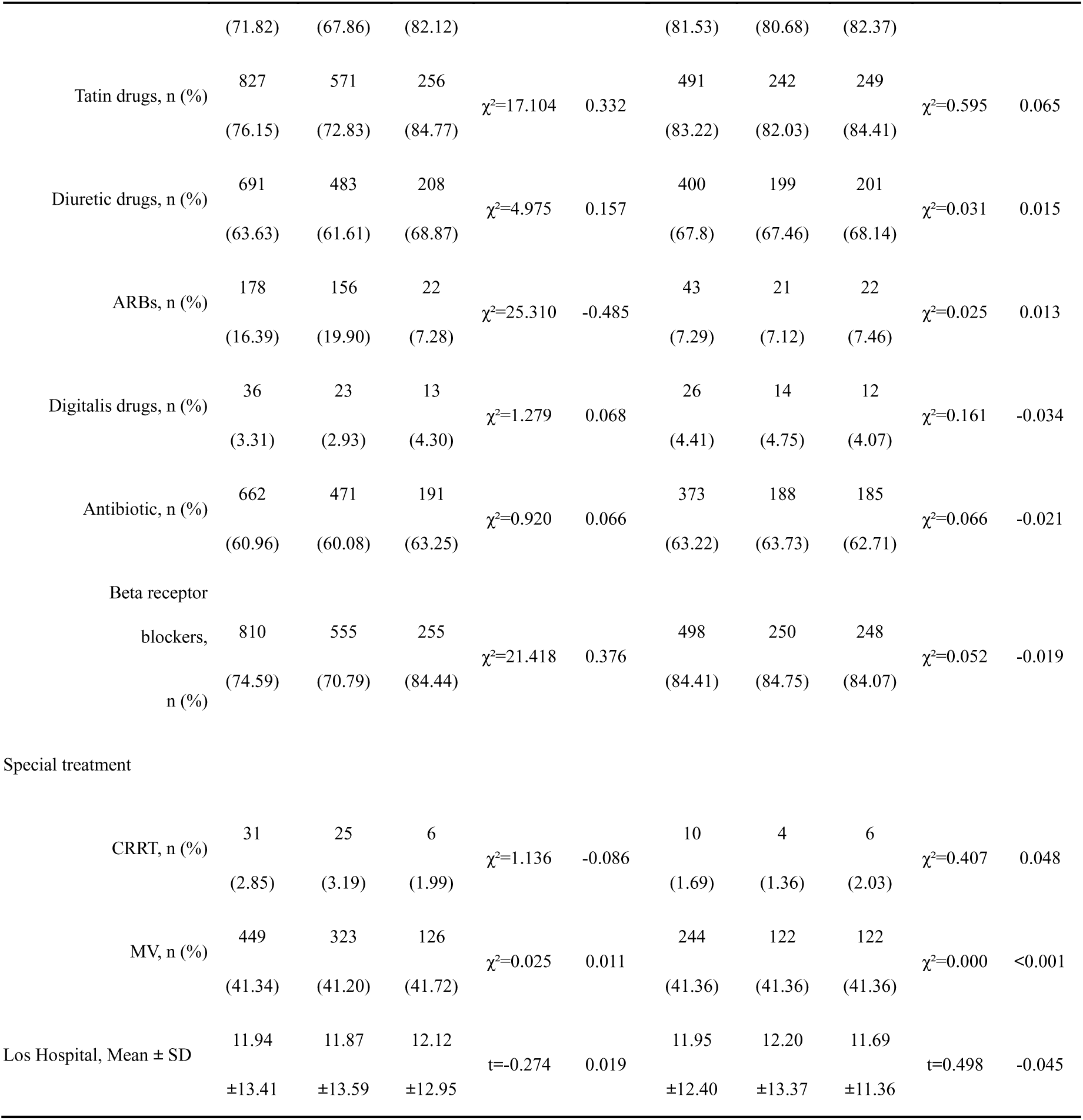
Comparison of baseline characteristics before and after PSM.

### Primary outcome

Figure 2 shows the Kaplan-Meier curve for 3-year all-cause mortality according to ACEI use. The analysis revealed that ACEI use significantly reduced 3-year all-cause mortality (HR, 0.55;95% CI, 0.41-0.74; P<0.001). Univariate Cox proportional hazards analysis showed that ACEI use lowered the risk of death within three years (HR, 0.55; 95% CI, 0.40-0.75; P<0.001). In addition, factors such as age, CCI, atrial fibrillation, congestive heart failure, chronic lung disease, kidney disease, malignant tumor, use of ARBs, and digitalis use all showed statistical significance. Multivariate Cox proportional hazards analysis adjusting for relevant factors showed that ACEI use continued to reduce the risk of 3-year all-cause death (HR, 0.59; 95% CI, 0.43-0.79; P<0.001), as presented in Table 2.

**Figure 2.**
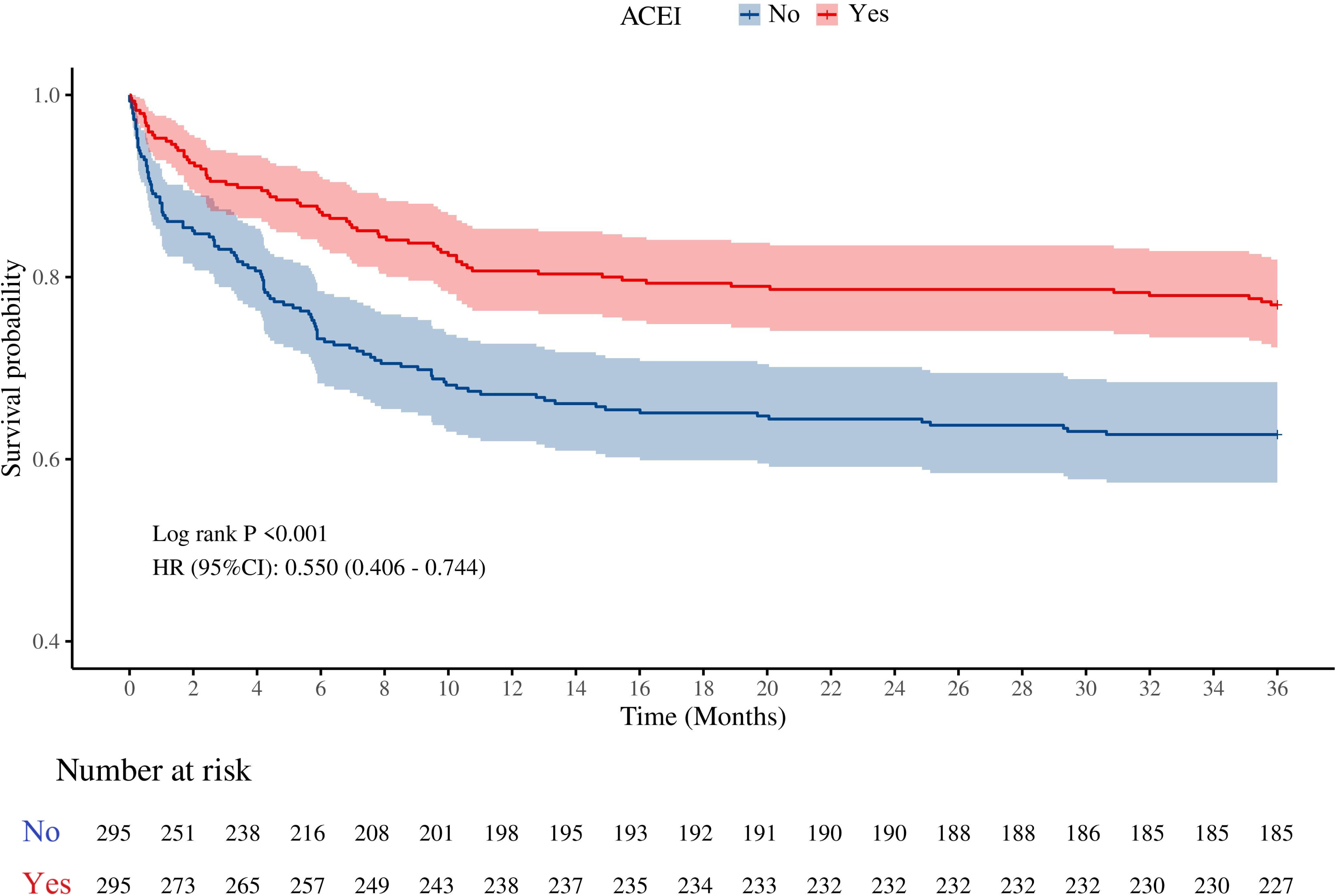
Kaplan-Meier curves for 3-year all-cause mortality matched by ACEI use

**Table 2.** Univariate and Multivariate Cox Models for 3-Year all-cause mortality.

| Variables | Univariate analysis |  |  |  |  | Multivariate analysis |  |  |  |  |
| --- | --- | --- | --- | --- | --- | --- | --- | --- | --- | --- |
| | $\beta$ | S.E | Z | P | HR (95%CI) | $\beta$ | S.E | Z | P | HR (95%CI) |
| Demographic data |  |  |  |  |  |  |  |  |  |  |
| Age | 0.05 | 0.01 | 7.03 | <0.001 | 1.05<br>(1.04 ~ 1.06) | 0.04 | 0.01 | 5.61 | <0.001 | 1.05<br>(1.03 ~ 1.06) |
| Male | -0.20 | 0.15 | -1.38 | 0.167 | 0.82<br>(0.61 ~ 1.09) |  |  |  |  |  |
| Comorbidity |  |  |  |  |  |  |  |  |  |  |
| CCI | 0.22 | 0.03 | 7.86 | <0.001 | 1.25<br>(1.18 ~ 1.32) | 0.16 | 0.03 | 5.50 | <0.001 | 1.18<br>(1.11 ~ 1.25) |
| OLD MI | 0.05 | 0.23 | 0.23 | 0.819 | 1.05<br>(0.67 ~ 1.67) |  |  |  |  |  |
| Atrial<br>Fibrillation | 0.32 | 0.16 | 2.06 | 0.039 | 1.38<br>(1.02 ~ 1.88) |  |  |  |  |  |
| Congestive<br>Heart Failure, n<br>(%) | 0.47 | 0.16 | 2.98 | 0.003 | 1.60<br>(1.18 ~ 2.19) |  |  |  |  |  |
| Cerebrovascul<br>ar Disease, n (%) | 0.08 | 0.20 | 0.40 | 0.686 | 1.08<br>(0.74 ~ 1.59) |  |  |  |  |  |
| Chronic<br>Pulmonary<br>Disease, n (%) | 0.35 | 0.16 | 2.12 | 0.034 | 1.42<br>(1.03 ~ 1.96) | 0.26 | 0.17 | 1.52 | 0.129 | 1.29<br>(0.93 ~ 1.80) |
| Diabetes<br>without cc, n (%) | -0.14 | 0.18 | -0.78 | 0.435 | 0.87<br>(0.61 ~ 1.23) |  |  |  |  |  |
| Diabetes with<br>cc, n (%) | 0.23 | 0.16 | 1.44 | 0.150 | 1.26<br>(0.92 ~ 1.73) |  |  |  |  |  |
| Renal<br>Disease, n (%) | 0.53 | 0.15 | 3.51 | <0.001 | 1.69<br>(1.26 ~ 2.27) |  |  |  |  |  |
| Malignant<br>Cancer, n (%) | 0.82 | 0.21 | 3.84 | <0.001 | 2.26<br>(1.49 ~ 3.43) |  |  |  |  |  |
| | $\beta$ | S.E | Z | P | HR (95%CI) | $\beta$ | S.E | Z | P | HR (95%CI) |
| Hypertension,<br>n (%) | -0.06 | 0.16 | -0.35 | 0.725 | 0.95<br>(0.69 ~ 1.29) |  |  |  |  |  |
| Hyperlipidemi<br>a, n (%) | -0.09 | 0.15 | -0.57 | 0.567 | 0.92<br>(0.68 ~ 1.23) |  |  |  |  |  |
| Risk factors for<br>T2MI |  |  |  |  |  |  |  |  |  |  |
| Sepsis | -0.03 | 0.20 | -0.16 | 0.873 | 0.97<br>(0.65 ~ 1.44) | -0.53 | 0.36 | -1.45 | 0.147 | 0.59<br>(0.29 ~ 1.20) |
| ARDS | -0.71 | 0.97 | -0.73 | 0.465 | 0.49<br>(0.07 ~ 3.30) |  |  |  |  |  |
| AKI | 0.28 | 0.16 | 1.81 | 0.070 | 1.33<br>(0.98 ~ 1.80) |  |  |  |  |  |
| Septic Shock | 0.11 | 0.23 | 0.50 | 0.619 | 1.12<br>(0.71 ~ 1.76) | 0.74 | 0.41 | 1.81 | 0.071 | 2.11<br>(0.94 ~ 4.73) |
| Tachycardia | 0.25 | 0.21 | 1.24 | 0.217 | 1.29<br>(0.86 ~ 1.93) | 0.46 | 0.22 | 2.11 | 0.035 | 1.58<br>(1.03 ~ 2.41) |
| Bradycardia | 0.27 | 0.37 | 0.73 | 0.463 | 1.32<br>(0.63 ~ 2.74) |  |  |  |  |  |
| ARF hypoxia | 0.18 | 0.17 | 1.06 | 0.290 | 1.19<br>(0.86 ~ 1.66) |  |  |  |  |  |
| Hypotension | -0.34 | 0.28 | -1.21 | 0.227 | 0.71<br>(0.41 ~ 1.24) |  |  |  |  |  |
| Acute<br>posthemorrhagic<br>anemia | -0.14 | 0.21 | -0.65 | 0.513 | 0.87<br>(0.58 ~ 1.32) |  |  |  |  |  |
| Medication status |  |  |  |  |  |  |  |  |  |  |
| Vasoactive agent | 0.10 | 0.15 | 0.68 | 0.498 | 1.11<br>(0.82 ~ 1.49) |  |  |  |  |  |
| ACEI | -0.60 | 0.16 | -3.77 | <0.001 | 0.55<br>(0.40 ~ 0.75) | -0.54 | 0.16 | -3.43 | <0.001 | 0.59<br>(0.43 ~ 0.79) |
| | $\beta$ | S.E | Z | P | HR (95%CI) | $\beta$ | S.E | Z | P | HR (95%CI) |
| Anticoagulant | 0.22 | 0.37 | 0.59 | 0.558 | 1.25<br>(0.60 ~ 2.60) |  |  |  |  |  |
| Antiplatelet | 0.02 | 0.20 | 0.09 | 0.928 | 1.02<br>(0.69 ~ 1.51) |  |  |  |  |  |
| Tatin drugs | 0.02 | 0.20 | 0.09 | 0.925 | 1.02<br>(0.69 ~ 1.49) |  |  |  |  |  |
| Diuretic drugs | 0.12 | 0.17 | 0.74 | 0.457 | 1.13<br>(0.82 ~ 1.58) |  |  |  |  |  |
| ARBs | -0.91 | 0.41 | -2.24 | 0.025 | 0.40<br>(0.18 ~ 0.89) | -0.78 | 0.42 | -1.84 | 0.066 | 0.46<br>(0.20 ~ 1.05) |
| Digitalis drugs | 0.65 | 0.33 | 1.99 | 0.046 | 1.92<br>(1.01 ~ 3.66) | 0.88 | 0.30 | 2.90 | 0.004 | 2.42<br>(1.33 ~ 4.40) |
| Antibiotic | 0.18 | 0.16 | 1.15 | 0.251 | 1.20<br>(0.88 ~ 1.64) |  |  |  |  |  |
| Beta receptor<br>blockers | -0.15 | 0.19 | -0.79 | 0.430 | 0.86<br>(0.60 ~ 1.25) |  |  |  |  |  |
| Special treatment |  |  |  |  |  |  |  |  |  |  |
| CRRT | -0.52 | 0.68 | -0.77 | 0.442 | 0.59<br>(0.16 ~ 2.25) |  |  |  |  |  |
| MV | -0.13 | 0.16 | -0.84 | 0.400 | 0.88<br>(0.64 ~ 1.19) |  |  |  |  |  |
| Los Hospital | 0.01 | 0.00 | 1.94 | 0.053 | 1.01<br>(1.00 ~ 1.02) | 0.01 | 0.01 | 2.67 | 0.008 | 1.01<br>(1.01 ~ 1.03) |

HR: Hazard Ratio, CI: Confidence Interval

### Subgroup analysis

Figure 3 shows the results of subgroup analysis of 3-year all-cause mortality in the matched cohort. The analysis found that in-hospital ACEI use significantly reduced the 3-year mortality risk in patients with T2MI across several subgroups. These subgroups included patients aged ≥80 years (HR, 0.59; 95% CI, 0.39-0.90; P=0.015) and 65-80 years (HR, 0.43; 95% CI, 0.25-0.74; P=0.002), those with CCI >8 (HR, 0.54; 95% CI, 0.35-0.83; P=0.005) or 7-8 (HR, 0.40; 95% CI, 0.21-0.75; P=0.004), patients without tachycardia (HR, 0.55; 95% CI, 0.40-0.77; P<0.001), and patients not using digitalis (HR, 0.58; 95% CI, 0.42-0.79; P<0.001).

**Figure 3.**
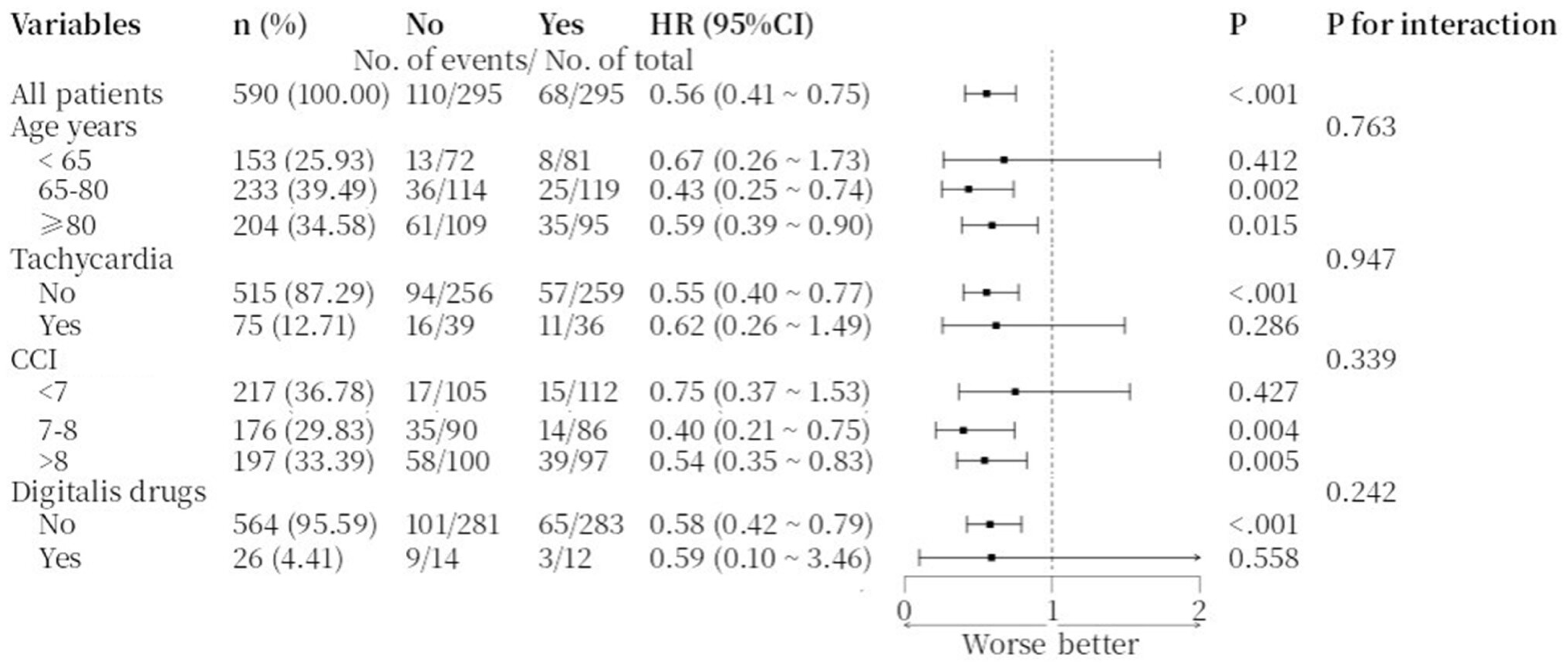
Subgroup analysis of 3-year all-cause mortality rates in the matched cohort

### Sensitivity analysis

The Cox regression analysis of multiple models showed that in-hospital ACEI use significantly reduced the 3-year all-cause mortality rate. This effect was consistent across all models: crude (HR 0.55, 95% CI 0.41-0.74; P<0.001, Model 1), adjusted for age and gender (HR 0.54, 95% CI 0.40-0.74; P<0.001, Model 2), further adjusted for medical history (HR 0.57, 95% CI 0.42-0.78; P<0.001, Model 3), and additionally adjusted for concomitant medications (HR 0.58, 95% CI 0.42-0.80; P<0.001, Model 4) (Table 3).

**Table 3.** Analysis of multiple Cox regression models on the association between ACEI use and 3-year all-cause mortality.

| Variables | Model1 |  | Model2 |  | Model3 |  | Model4 |  |
| --- | --- | --- | --- | --- | --- | --- | --- | --- |
|  | HR (95%CI) | P | HR (95%CI) | P | HR (95%CI) | P | HR (95%CI) | P |
| ACEI |  |  |  |  |  |  |  |  |
| No | 1.00 (Reference) |  | 1.00 (Reference) |  | 1.00 (Reference) |  | 1.00 (Reference) |  |
| Yes | 0.55<br>(0.41 ~ 0.74) | <0.001 | 0.54<br>(0.40 ~ 0.74) | <0.001 | 0.57<br>(0.42 ~ 0.78) | <0.001 | 0.58<br>(0.42 ~ 0.80) | <0.001 |

HR: Hazard Ratio, CI: Confidence Interval
Model1: Crude
Model2: Adjust: gender, age
Model3: Adjust: gender, OLD\_MI, Atrial\_fibrillation, congestive\_heart\_failure, cerebrovascular\_disease, chronic\_pulmonary\_disease, diabetes\_without\_cc, diabetes\_with\_cc, renal\_disease, malignant\_cancer, hypertension, hyperlipidemia, age, CCI
Model4: Adjust: gender, OLD\_MI, Atrial\_fibrillation, Anticoagulant, Diuretic\_drugs, ARBs, Digitalis\_drugs, antibiotic, Beta\_receptor\_blockers, Antiplatelet, Tatin\_drugs, congestive\_heart\_failure, cerebrovascular\_disease, chronic\_pulmonary\_disease, diabetes\_without\_cc, diabetes\_with\_cc, renal\_disease, malignant\_cancer, hypertension, hyperlipidemia, Vasoactive\_agent, age, CCI

### Secondary outcome

#### 180-day mortality

Figure 4 presents the Kaplan-Meier curve for 180-day all-cause mortality categorized by ACEI use. It reveals that ACEI significantly reduces mortality (HR, 0.43; 95% CI, 0.29-0.64; P<0.001). Univariate Cox analysis indicates that ACEI use reduces the risk of 180-day all-cause mortality in patients (HR, 0.43; 95% CI, 0.29-0.65; P<0.001). In addition, factors such as age, CCI, atrial fibrillation, chronic lung disease, malignant tumor, ARDS, use of digitalis, and CRRT all showed statistical significance. Multivariate Cox analysis adjusting for relevant factors showed that ACEI use remained significantly associated with lower 180-day all-cause mortality (HR, 0.49; 95% CI, 0.33-0.72; P<0.001) (Supplementary table 3).

**Figure 4.**
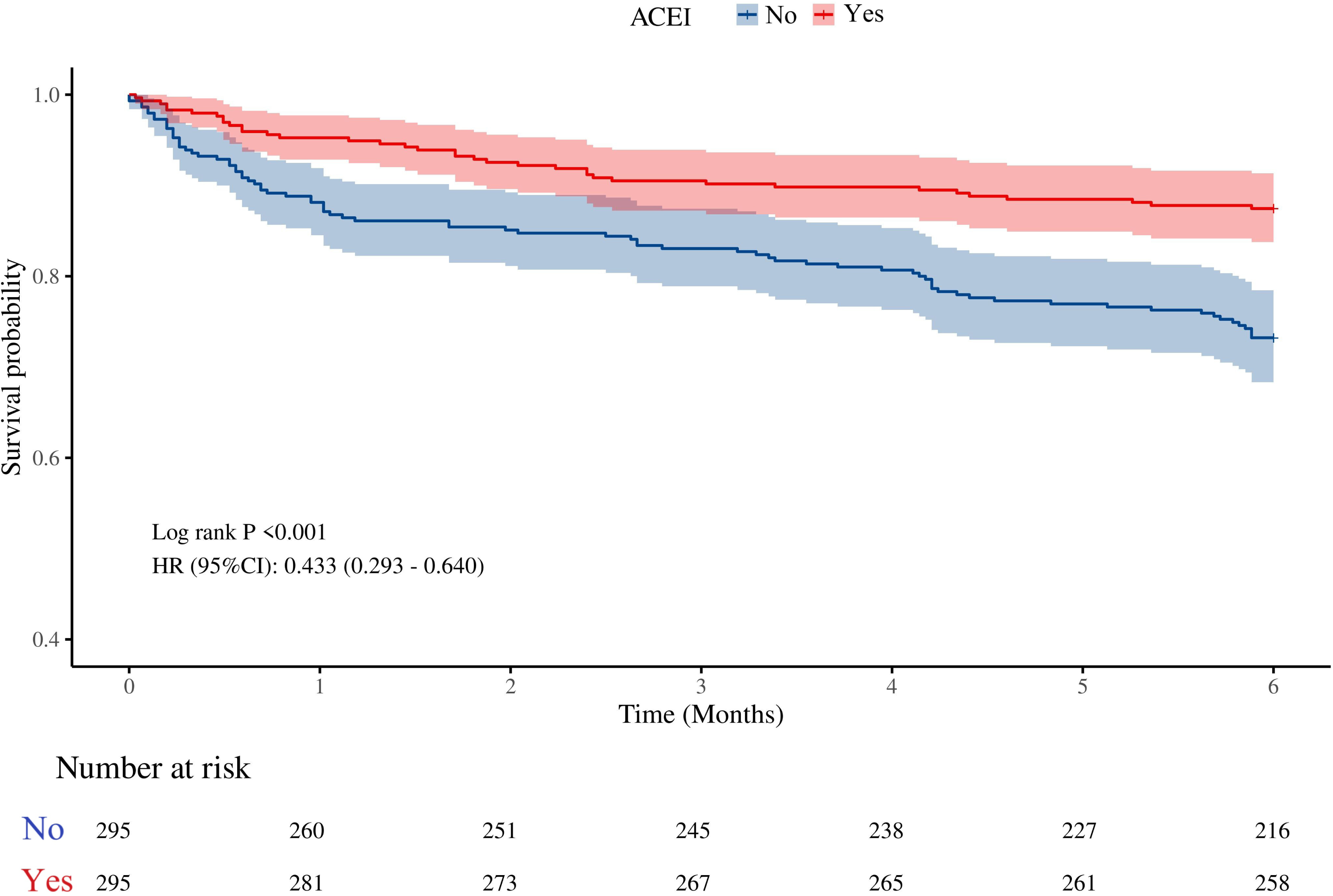
Kaplan-Meier curves for 180-day all-cause mortality matched by ACEI use

Sensitivity analysis using a Cox regression multi-model approach showed that crude 180-day all-cause mortality was significantly lower in the ACEI group (HR, 0.43; 95% CI, 0.29-0.64; P<0.001, Model 1, Supplementary table 4). After adjusting for age and gender (Model 2), in-hospital ACEI use remained significantly associated with reduced mortality (HR, 0.44; 95% CI, 0.30-0.65; P<0.001). Further adjustments for previous relevant medical history (Model 3) and relevant medications (Model 4) also demonstrated significant mortality reductions (HR, 0.47 and 0.49, respectively; both P<0.001).

#### 3-year all-cause readmission rate and T2MI recurrence rate

The Kaplan-Meier curve of the 3-year all-cause readmission rate was plotted based on ACEI use. As shown in Figure 5(a), ACEI use had no effect on the 3-year all-cause readmission risk (HR, 1.003; 95% CI, 0.776-1.295; P=0.984). Univariate Cox proportional hazards model analysis indicated that ACEI use had no effect on the 3-year all-cause readmission risk (HR, 0.89; 95% CI, 0.65-1.20; P=0.441). However, factors such as congestive heart failure, diabetes mellitus, kidney disease, acute kidney injury, and digitalis use all showed statistical significance. Multivariate Cox proportional hazards model analysis adjusted for relevant factors confirmed that ACEI use still had no effect on the 3-year all-cause readmission risk (HR, 1.00; 95% CI, 1.00-1.00; P=0.664) (Supplementary Table 5).

**Figure 5.**
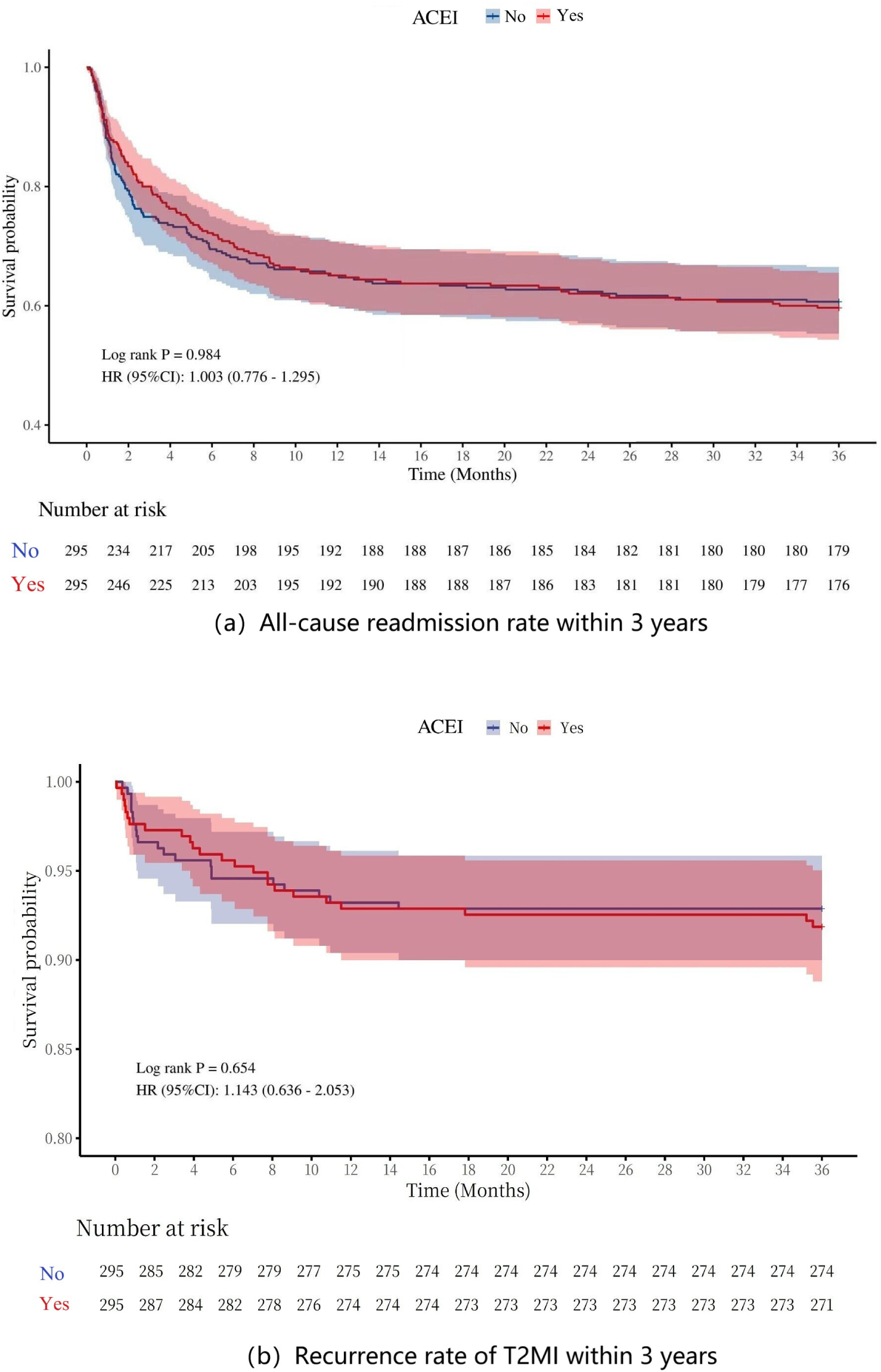
Kaplan-Meier curves for readmission and recurrence rates within 3 years of T2MI. (a): All-cause readmission rate; (b): T2MI recurrence rate

Figure 5(b) shows the Kaplan-Meier curve for the 3-year T2MI recurrence rate. It reveals that ACEI use has no significant effect on recurrence risk (HR, 1.14; 95% CI, 0.64-2.05; P=0.654). Univariate Cox proportional hazards analysis indicated that ACEI use had no effect on the risk of 3-year T2MI recurrence (HR, 1.14; 95% CI, 0.64-2.04; P=0.652). By contrast, factors such as congestive heart failure, diabetes mellitus, kidney disease, acute kidney injury, and digitalis use showed statistical significance. Further multivariate Cox proportional hazards analysis, adjusted for relevant factors, confirmed that ACEI use still had no effect on 3-year T2MI recurrence risk (HR, 1.10; 95% CI, 0.59-2.04; P=0.762) (Supplementary Table 6).

## Discussion

This is the first study to investigate the relationship between ACEI use and prognosis in T2MI patients discharged alive. We found that in-hospital ACEI use significantly reduced both short-term (180 days) and long-term (3 years) all-cause mortality rates, but did not affect post-discharge readmission or T2MI recurrence. The study findings provide real-world evidence supporting ACEI use in T2MI patients and may guide the development of treatment strategies.

ACEI dilates blood vessels, reduces cardiac load, and delays myocardial remodeling. It achieves these effects by inhibiting the RAS, reducing Ang II production, and promoting bradykinin accumulation [15–17]. The cardioprotective effect of ACEI in T1MI has been well established. Existing studies have found that AMI patients treated with ACEI have improved 5-year survival rates compared to those without ACEI treatment [15]. ACEI treatment reduces all-cause mortality in AMI patients undergoing percutaneous coronary intervention [22]. Additionally, after 2 years of follow-up, patients treated with ACEI show lower mortality rates than those treated with ARBs [15]. Moreover, during hospitalization of AMI patients, those who do not adhere to ACEI treatment have significantly higher mortality rates than those who do, regardless of statin or beta-blocker use [17]. These findings collectively suggest that ACEI provides clear benefits to AMI patients. Nevertheless, because of the different underlying mechanisms between T2MI and T1MI, as well as the influence of comorbidities, it remains unclear whether ACEI treatment benefits T2MI patients. Further research is needed to clarify the benefits of ACEI treatment in T2MI patients.

A recent study found that in-hospital use of ACEI may improve both in-hospital and 2-year survival rates of T2MI patients [19]. This study further provides supporting evidence for this association. This study found that among T2MI patients discharged alive, in-hospital use of ACEI significantly reduces all-cause mortality after discharge. This mortality reduction was observed in both the short-term and long-term follow-up periods. This effect may be explained by two mechanisms. First, ACEI reduces infarct size and inflammatory response. After myocardial infarction, the renin-angiotensin-aldosterone system (RAAS) activates, with Ang II as its key hormone [23]. Its expression level increases within hours and can persist for several days [24]. This leads to enhanced coronary vasoconstriction, which reduces oxygen supply while increasing oxygen consumption. It also mediates inflammatory factors, exacerbating myocardial ischemic damage [25–27]. In addition, infarct size is positively correlated with cardiac angiotensin-converting enzyme(ACE) activity [28]. Therefore, ACEI can reduce the infarct size by inhibiting the production of Ang II [29] and suppress the inflammatory response [30], thereby improving cardiac function as early as possible in the short term. 2) The use of ACEI helps preserve cardiac structure and prevent adverse remodeling. Studies have shown that in patients with first anterior wall myocardial infarction, administering captopril within 24 hours after symptom onset for 14 days significantly reduced the incidence of ventricular dilatation [31]. Additionally, it could attenuate the early stage of left ventricular remodeling [32]. In addition, reduced baroreflex sensitivity is closely associated with the occurrence of ventricular tachycardia. It also increases the risk of hemodynamic deterioration during tachycardia [33] and is linked to higher cardiac mortality [34, 35]. However, treatment with captopril within 4 days after symptom onset in myocardial infarction patients can significantly increase baroreflex sensitivity [36], with an acute enhancing effect observable as early as 1 hour after administration [37], thereby reducing the risk of poor prognosis.

However, this study found that ACEI did not reduce either the all-cause readmission rate or the recurrence rate of T2MI. This result is inconsistent with previous research on the effect of ACEI on myocardial infarction. One possible reason is that non-coronary events—such as anemia, hypoxemia, hypotension, and other major risk factors for T2MI [2, 38]—raise the risk of patient readmission [39]. However, ACEI mainly acts on the cardiovascular system [40]. Therefore, it cannot significantly reduce the risk of readmission or recurrence caused by non-cardiovascular events.

In addition, existing studie has found [41] that ACEI-based treatment regimens provide greater cardio-cerebrovascular protection to elderly patients. This protection is stronger compared with that observed in younger patients. This includes a significant reduction in the risks of stroke, death, and heart failure. The subgroup analysis further confirms that elderly patients benefit more from ACEI treatment. For T2MI patients aged ≥65 years, ACEI may provide greater benefits when combined with active anti-infection therapy, anemia correction, and other treatments targeting the primary disease. Additionally, the study found that ACEI use benefits patients without tachyarrhythmia, with a CCI score ≥7, and not using digitalis. Therefore, ACEI should be prioritized for this patient population in clinical practice.

This study has several limitations. First, it is a single-center retrospective study with a relatively small sample size. Further verification through multi-center prospective studies is still needed. Second, the study only included data on in-hospital ACEI use and lacked information on continued drug use outside the hospital, which might underestimate the actual impact of long-term medication. Third, some hospitalized patients did not receive ACEI due to contraindications or adverse reactions, potentially leading to selection bias. Fourth, data on the specific dosage and course of ACEI are missing, which makes it impossible to analyze the dose-effect relationship and affects the inference of the optimal medication regimen.

## Conclusion

This study found that using ACEI during hospitalization significantly reduces both short- and long-term all-cause mortality in T2MI patients discharged alive, while it did not affect patients’ readmission rates or T2MI recurrence rates. This finding expands the clinical applications of ACEI in populations with myocardial infarction caused by different mechanisms. Nevertheless, due to study design limitations, clinicians must carefully evaluate the indications and contraindications for each patient. Future prospective studies should further clarify the optimal dosage, timing, and patient selection for ACEI administration.<colcnt=2>

## List of abbreviations

ACEI: Angiotensin converting enzyme inhibitor
T2MI: Type 2 myocardial infarction
MIMIC-IV: Medical Information Mart for Intensive Care IV
PSM: Propensity score matching
SGLT2: Sodium-dependent glucose transporters 2
GLP-1: Glucagon-like peptide-1
T1MI: Type 1 myocardial infarction
RAS: Renin-angiotensin system
LVEF: Left ventricular ejection fraction
MIMIC-IV 3.1: Medical Information Mart for Intensive Care IV version 3.1
ICD: International classification of diseases
SQL: Structured Query Language
CCI: Charlson Comorbidity Index
ARDS: Acute respiratory distress syndrome
ARBs: Angiotensin II receptor blockers
CRRT: Continuous renal replacement therapy
HR: Hazard ratio
CI: Confidence interval
SMD: Standardized mean difference
VIF: Variance inflation factor
Ang Ⅱ: Angiotensin II
AMI: Acute myocardial infarction
RAAS: Renin-angiotensin-aldosterone system
ACE: Angiotensin-converting enzyme
MV: Mechanical ventilation

## Declarations

### Ethics approval and consent to participate

The study protocol was in accordance with the Declaration of Helsinki and was reviewed and approved by the Ethics Committee of Shanghai Jiading District Central Hospital (2025-34), an affiliated teaching hospital of Shanghai University of Medicine & Health Sciences. In accordance with national laws and institutional requirements, written informed consent was not required of the participants because of the nature of the study.

### Authors’ contributions

ZJY, LJN and FWY: carried out the studies, participated in collecting data, and drafted the manuscript. LY, WYX, MXY: participated in collecting data and helped to draft the manuscript. LYY and LWL: performed the statistical analysis. WF and JXF: design, review and editing the manuscript. All authors read and approved the final manuscript.

## Acknowledgements

Not applicable

## Funding

The study was supported by Shanghai Municipal Health Commission Health Industry Clinical Research Project (202340100), Jiading District Central Hospital Clinical Research Support Program (2025LY14 and 2025LY18). The funding sources had no role in the design of the study; in the collection, analysis, and interpretation of data; or in the writing of the manuscript.

### Data availability statement

The data from public databases were available on the MIMIC-IV website at https://mimic.physionet.org/.

### Consent for publication

Not applicable.

---

**Supplementary Figure 1** SMD before and after PSM

**Supplementary Figure 2** Distribution balance before and after PSM

## References

1. White K, Kinarivala M, Scott I. Diagnostic features, management and prognosis of type 2 myocardial infarction compared to type 1 myocardial infarction: a systematic review and meta-analysis. BMJ Open. 2022;12(2):e055755.

2. Coscia T, Nestelberger T, Boeddinghaus J, Lopez-Ayala P, Koechlin L, Miró Ò, et al. Characteristics and Outcomes of Type 2 Myocardial Infarction. JAMA Cardiol. 2022;7(4):427–34.

3. Thygesen K, Alpert JS, Jaffe AS, Chaitman BR, Bax JJ, Morrow DA, et al. Fourth Universal Definition of Myocardial Infarction (2018). J Am Coll Cardiol. 2018;72(18):2231–64.

4. White HD, Steg PG, Szarek M, Bhatt DL, Bittner VA, Diaz R, et al. Effects of alirocumab on types of myocardial infarction: insights from the ODYSSEY OUTCOMES trial. Eur Heart J. 2019;40(33):2801–9.

5. Lambrakis K, French JK, Scott IA, Briffa T, Brieger D, Farkouh ME, et al. The appropriateness of coronary investigation in myocardial injury and type 2 myocardial infarction (ACT-2): A randomized trial design. Am Heart J. 2019;208:11–20.

6. Fitchett D, Zinman B, Inzucchi SE, Wanner C, Anker SD, Pocock S, et al. Effect of empagliflozin on total myocardial infarction events by type and additional coronary outcomes: insights from the randomized EMPA-REG OUTCOME trial. Cardiovasc Diabetol. 2024;23(1):248.

7. Krychtiuk KA, Marquis-Gravel G, Murphy S, Alexander KP, Chiswell K, Green JB, et al. Effects of albiglutide on myocardial infarction and ischaemic heart disease outcomes in patients with type 2 diabetes and cardiovascular disease in the Harmony Outcomes trial. Eur Heart J Cardiovasc Pharmacother. 2024;10(4):279–88.

8. Guimarães PO, Leonardi S, Huang Z, Wallentin L, de Werf FV, Aylward PE, et al. Clinical features and outcomes of patients with type 2 myocardial infarction: insights from the Thrombin Receptor Antagonist for Clinical Event Reduction in Acute Coronary Syndrome (TRACER) trial. Am Heart J 2018;196:28–35. 10.1016/j.ahj.2017.10.007.

9. Shah AS, McAllister DA, Mills R, Lee KK, Churchhouse AM, Fleming KM, et al. Sensitive troponin assay and the classification of myocardial infarction. Am J Med. 2015;128(5):493–501.e3.

10. Arora S, Strassle PD, Qamar A, Wheeler EN, Levine AL, Misenheimer JA, et al. Impact of Type 2 Myocardial Infarction (MI) on Hospital-Level MI Outcomes: Implications for Quality and Public Reporting. J Am Heart Assoc. 2018;7(7).

11. Sandoval Y, Smith SW, Sexter A, Thordsen SE, Bruen CA, Carlson MD, et al. Type 1 and 2 Myocardial Infarction and Myocardial Injury: Clinical Transition to High-Sensitivity Cardiac Troponin I. Am J Med. 2017;130(12):1431–9.e4.

12. Singh A, Gupta A, DeFilippis EM, Qamar A, Biery DW, Almarzooq Z, et al. Cardiovascular Mortality After Type 1 and Type 2 Myocardial Infarction in Young Adults. J Am Coll Cardiol. 2020;75(9):1003–13.

13. Kimenai DM, Lindahl B, Chapman AR, Baron T, Gard A, Wereski R, et al. Sex differences in investigations and outcomes among patients with type 2 myocardial infarction. Heart. 2021;107(18):1480–6.

14. Eggers KM, Baron T, Gard A, Lindahl B. Clinical and prognostic implications of high-sensitivity cardiac troponin T concentrations in type 2 non-ST elevation myocardial infarction. Int J Cardiol Heart Vasc. 2022;39:100972.

15. Hara M, Sakata Y, Nakatani D, Suna S, Usami M, Matsumoto S, et al. Comparison of 5-year survival after acute myocardial infarction using angiotensin-converting enzyme inhibitor versus angiotensin II receptor blocker. Am J Cardiol. 2014;114(1):1–8.

16. Korhonen MJ, Robinson JG, Annis IE, Hickson RP, Bell JS, Hartikainen J, et al. Adherence Tradeoff to Multiple Preventive Therapies and All-Cause Mortality After Acute Myocardial Infarction. J Am Coll Cardiol. 2017;70(13):1543–54.

17. Ahn JH, Hyun JY, Jeong MH, Kim JH, Hong YJ, Sim DS, et al. Comparative effect of angiotensin converting enzyme inhibitor versus angiotensin ii type i receptor blocker in acute myocardial infarction with non-obstructive coronary arteries; from the Korea Acute Myocardial Infarction Registry - National Institute of Health. Cardiol J. 2021;28(5):738–45.

18. Escobar J, Rawat A, Maradiaga F, Isaak AK, Zainab S, Arusi Dari M, et al. Comparison of Outcomes Between Angiotensin-Converting Enzyme Inhibitors and Angiotensin II Receptor Blockers in Patients With Myocardial Infarction: A Meta-Analysis. Cureus. 2023;15(10):e47954.

19. Šerpytis R, Lizaitis M, Majauskienė E, Navickas P, Glaveckaitė S, Petrulionienė Ž, et al. Type 2 Myocardial Infarction and Long-Term Mortality Risk Factors: A Retrospective Cohort Study. Adv Ther. 2023;40(5):2471–80.

20. Johnson AEW, Bulgarelli L, Shen L, Gayles A, Shammout A, Horng S, et al. MIMIC-IV, a freely accessible electronic health record dataset. Sci Data. 2023;10(1):1.

21. Goldberger AL, Amaral LA, Glass L, Hausdorff JM, Ivanov PC, Mark RG, et al. PhysioBank, PhysioToolkit, and PhysioNet: components of a new research resource for complex physiologic signals. Circulation. 2000;101(23):E215–20.

22. Savarese G, Costanzo P, Cleland JG, Vassallo E, Ruggiero D, Rosano G, et al. A meta-analysis reporting effects of angiotensin-converting enzyme inhibitors and angiotensin receptor blockers in patients without heart failure. J Am Coll Cardiol. 2013;61(2):131–42.

23. Jalowy A, Schulz R, Heusch G. AT1 receptor blockade in experimental myocardial ischemia/reperfusion. J Am Soc Nephrol. 1999;10 Suppl 11:S129–36.

24. Oyamada S, Bianchi C, Takai S, Robich MP, Clements RT, Chu L, Sellke FW. Impact of acute myocardial ischemia reperfusion on the tissue and blood-borne renin-angiotensin system. Basic Res Cardiol. 2010 Jul;105(4):513–22. doi: 10.1007/s00395-010-0093-4. Epub 2010 Mar 26. PMID: 20340028.

25. Matus M, Kucerova D, Kruzliak P, Adameova A, Doka G, Turcekova K, Kmecova J, Kyselovic J, Krenek P, Kirchhefer U, Mueller FU, Boknik P, Klimas J. Upregulation of SERCA2a following short-term ACE inhibition (by enalaprilat) alters contractile performance and arrhythmogenicity of healthy myocardium in rat. Mol Cell Biochem. 2015 May;403(1-2):199–208. doi: 10.1007/s11010-015-2350-1. Epub 2015 Feb 8. PMID: 25663023.

26. Haymann JP, Hammoudi N, Stankovic Stojanovic K, Galacteros F, Habibi A, Avellino V, Bartolucci P, Benzerara Y, Arlet JB, Djebbar M, Letavernier E, Grateau G, Tabibzadeh N, Girshovich A, Chaignon M, Girot R, Levy P, Lionnet F. Renin-angiotensin system blockade promotes a cardio-renal protection in albuminuric homozygous sickle cell patients. Br J Haematol. 2017 Dec;179(5):820–828. doi: 10.1111/bjh.14969. Epub 2017 Oct 19. PMID: 29048108.

27. Dai W, Kloner RA. Potential role of renin-angiotensin system blockade for preventing myocardial ischemia/reperfusion injury and remodeling after myocardial infarction. Postgrad Med. 2011;123(2):49–55.

28. McCarthy CP, Kolte D, Kennedy KF, Vaduganathan M, Wasfy JH, Januzzi JL Jr. Patient Characteristics and Clinical Outcomes of Type 1 Versus Type 2 Myocardial Infarction. J Am Coll Cardiol. 2021 Feb 23;77(7):848–857. doi: 10.1016/j.jacc.2020.12.034. PMID: 33602466.

29. Chen X, Minatoguchi S, Wang N, Arai M, Lu C, Uno Y, et al. Quinaprilat reduces myocardial infarct size involving nitric oxide production and mitochondrial KATP channel in rabbits. J Cardiovasc Pharmacol. 2003;41(6):938–45.

30. Sandmann S, Li J, Fritzenkötter C, Spormann J, Tiede K, Fischer JW, et al. Differential effects of olmesartan and ramipril on inflammatory response after myocardial infarction in rats. Blood Press. 2006;15(2):116–28.

31. Lambrecht S, Sarkisian L, Saaby L, Poulsen TS, Gerke O, Hosbond S, Diederichsen ACP, Thygesen K, Mickley H. Different causes of death in patients with myocardial infarction type 1, type 2, and myocardial injury. Am J Med 2018;131:548– 554.

32. Maraey A, Elzanaty AM, Salem M, Khalil M, Elsharnoby H, Younes A, Elsharnouby M, Nazir S, Elgendy IY, Siragy HM. Relation of Type 2 Myocardial Infarction and Readmission With Type 1 Myocardial Infarction in Hypertensive Crises (from a Nationwide Analysis). Am J Cardiol. 2021 Dec 15;161:56–62. doi: 10.1016/j.amjcard.2021.08.060. Erratum in: Am J Cardiol. 2022 Sep 1;178:180. doi: 10.1016/j.amjcard.2022.06.025. PMID: 34794619.

34. Lee CJ, Choi B, Pak H, Park JM, Lee JH, Lee SH. Genetic Variants Associated with Adverse Events after Angiotensin-Converting Enzyme Inhibitor Use: Replication after GWAS-Based Discovery. Yonsei Med J. 2022 Apr;63(4):342–348. doi: 10.3349/ymj.2022.63.4.342. PMID: 35352885; PMCID: PMC8965428.

35. Schmidt M, Mansfield KE, Bhaskaran K, et al. Serum creatinine elevation after renin-angiotensin system blockade and long term cardiorenal risks: Cohort study. BMJ 2017;356:j791.

36. Fang G, Annis IE, Farley JF, Mahendraratnam N, Hickson RP, Stürmer T, Robinson JG. Incidence of and Risk Factors for Severe Adverse Events in Elderly Patients Taking Angiotensin-Converting Enzyme Inhibitors or Angiotensin II Receptor Blockers after an Acute Myocardial Infarction. Pharmacotherapy. 2018 Jan;38(1):29–41. doi: 10.1002/phar.2051. Epub 2017 Dec 11. PMID: 29059475; PMCID: PMC5766359.

37. Marakas SA, Kyriakidis MK, Vourlioti AN, Petropoulakis PN, Toutouzas PK. Acute effect of captopril administration on baroreflex sensitivity in patients with acute myocardial infarction. Eur Heart J. 1995;16(7):914–21.

38. Wang F, Wu X, Hu SY, Wu YW, Ding Y, Ye LZ, et al. Type 2 myocardial infarction among critically ill elderly patients in the Intensive Care Unit: the clinical features and in-hospital prognosis. Aging Clin Exp Res. 2020;32(9):1801–7.

39. Rocheleau S, Eng-Frost J, Lambrakis K, Khan E, Chiang B, Wattchow N, et al. Twelve-Month Outcomes of Patients With Myocardial Injury not Due to Type-1 Myocardial Infarction. Heart Lung Circ. 2023;32(8):978–85.

40. Pavo N, Wurm R, Goliasch G, et al. Renin-angiotensin system fingerprints of heart failure with reduced ejection fraction. J Am Coll Cardiol. 2016;68(25):2912– 2914. doi: 10.1016/j.jacc.2016.10.017.

41. Liu X, Xie Z, Zhang Y, Huang J, Kuang L, Li X, Li H, Zou Y, Xiang T, Yin N, Zhou X, Yu J. Machine learning for predicting in-hospital mortality in elderly patients with heart failure combined with hypertension: a multicenter retrospective study. Cardiovasc Diabetol. 2024 Nov 15;23(1):407. doi: 10.1186/s12933-024-02503-9.

